# Patterns of reporting of influenza and pertussis vaccination during pregnancy to the Australian Immunisation Register, 2022

**DOI:** 10.1101/2025.01.23.25320512

**Authors:** Nicole Sonneveld, Elizabeth Wilson, Sonya Ennis, Jocelynne McRae, Kristine Macartney, Bette Liu

## Abstract

**Objectives:** Timely, accurate reporting of pregnancy vaccination coverage is key to evaluating pregnancy immunisation programs. We compared influenza and pertussis vaccination reporting on the New South Wales (NSW) Perinatal Data Collection (PDC) and the Australian Immunisation Register (AIR), to understand coverage, reporting, timing of vaccination and provider type.

**Design:** Retrospective population-based cohort study.

**Setting:** People giving birth at ≥20-weeks gestation (2017–2022) in the NSW PDC, linked to AIR-reported influenza and pertussis vaccinations.

**Main outcome measures:** Influenza and pertussis coverage according to 1) the PDC and 2) linked AIR records, and agreement between data sources by year (influenza: 2021–2022, pertussis: 2017–2022) and selected characteristics (2022). Description of provider type and timing for vaccinations reported to the AIR (2022).

**Results:** PDC-reported influenza and pertussis coverage peaked in 2020 (influenza: 58.8% (52,940/90,091), pertussis: 79.0% (71,169/90,091)), decreasing thereafter (influenza: 49.1% (43,744/89,112), pertussis: 77.6% (69,129/89,112) in 2022). Influenza and pertussis vaccination coverage as reported to the AIR increased from 34.7% (33,611/96,789) in 2021 to 44.8% (39,935/89,112) in 2022 and 24.4% (22,857/93,582) in 2017 to 51.6% (46,009/89,112) in 2022, respectively. Agreement between AIR and PDC improved over time, but remained suboptimal (influenza: 75.5%, pertussis: 56.6% in 2022). AIR and PDC agreement differed most by antenatal care model, particularly for pertussis vaccinations. In 2022, of pregnancy vaccinations reported to the AIR, >75% were provided in general practices. Co-administration occurred infrequently (8.8%).

**Conclusions:** Despite mandated reporting to the AIR, there was significant underreporting of pregnancy vaccinations to the AIR as compared with the NSW PDC. National collection of accurate and timely data on pregnancy vaccination coverage is needed to monitor vaccine uptake, and identify and monitor strategies to improve uptake, particularly with the new pregnancy RSV vaccine program in 2025. We identified co-administration as a potential strategy to improve influenza vaccination coverage during pregnancy.

## Introduction

Influenza and pertussis vaccination is recommended and funded under the Australian National Immunisation Program (NIP) during each pregnancy (1). Vaccination during pregnancy (henceforth: pregnancy vaccination) protects the pregnant person and their infant against severe illness (2-6). Influenza pregnancy vaccination was included on the NIP in 2010 (7); and all jurisdictions had implemented a pertussis pregnancy vaccination program by 2015, with a nationally-funded program commencing in 2018 (8). Timely, high-quality data on pregnancy vaccination coverage is required to accurately monitor and improve the impact of these programs, but national coverage estimates are unavailable (9). Population-level estimates based on jurisdictional Perinatal Data Collections (PDCs) have been reported (9- 13), but this occurs inconsistently across the country.

In NSW, the PDC captures fact of influenza and pertussis pregnancy vaccination but not the date or setting. Program-relevant information, such as gestation at vaccination or vaccine co-administration, cannot be determined based on the PDC. The Australian Immunisation Register (AIR) captures all vaccines given to people in Australia. While reporting of influenza and all NIP-funded vaccinations to the AIR became mandatory in 2021 (14), completeness of reporting is unknown. Information available on the AIR includes vaccination date and provider type. Until recently, the AIR did not capture pregnancy status at vaccination. While it is now possible to report whether vaccination occurred during pregnancy, vaccination coverage cannot easily be estimated based on AIR data alone, as the denominator (i.e. total number of pregnant people) is unavailable within the AIR.

Linking the PDC to the AIR allows us to establish if a person was pregnant when vaccinated, and to assess timing of and settings in which vaccines were provided. For vaccinations not captured in the PDC, such as COVID-19 and RSV vaccination (15), linkage of the PDC and AIR is also the most comprehensive method through which to estimate coverage.

This study had three aims: 1. To provide influenza and pertussis pregnancy vaccination coverage estimates based on the AIR and PDC, separately; 2. To describe timing and setting of vaccination encounters in pregnancy reported to the AIR, and 3. To assess the agreement of influenza and pertussis vaccination status reported on the AIR and PDC.

## Material and Methods

### Data Sources and Study Population

The NSW PDC is a population-based surveillance system that includes demographics for pregnant people and their infants, and pregnancy, labour, birth and perinatal outcomes. It covers all livebirths, and stillbirths of at least 20-weeks gestational age or 400-grams birthweight (16). Influenza and pertussis pregnancy vaccination status is assessed based on provider or patient-held record or verbal report (method of verification not captured). While some providers may verify vaccination status in the AIR, the PDC is not populated by information captured in the AIR.

The AIR is an Australian Government Department of Health and Aged Care national register of all vaccines given to people in Australia (17), to which immunisation providers can report using various methodologies. Reporting of influenza vaccination to the AIR was mandated on 1 March 2021; reporting of all NIP-funded vaccines was mandated on 1 July 2021 (14).

We performed a retrospective cohort study including all pregnancies reported to the NSW PDC that ended between 1 January 2017 and 31 December 2022 at ≥20-weeks gestation in people aged 12–55 years. People could be included multiple times if they carried multiple pregnancies to ≥20-weeks gestation in this period. Pregnancies were excluded if the pregnant person’s date of birth or gestation at the end of pregnancy was unavailable. From the NSW PDC, we obtained date of pregnant person’s birth, area of residence, main model of antenatal care, date of infant’s birth and gestation at the end of pregnancy, as well as influenza and pertussis pregnancy vaccination status (vaccinated yes/no/unknown).

The Centre for Health Record Linkage (CHeReL) linked the AIR to the PDC using record linkage methods (18,19). We included all influenza vaccines reported from 1 January 2021 (records prior to this date were not available for linkage) and all pertussis vaccines reported from 1 January 2016 in our study population. We obtained vaccination date, vaccine brand and vaccination provider type.

We defined PDC-reported vaccination coverage as the proportion of pregnancies during which receipt of vaccination was indicated on the PDC. We defined AIR-reported coverage as the proportion of pregnancies during which at least one influenza or pertussis-containing vaccine dose was reported to the AIR between the estimated last menstrual period (LMP; end of pregnancy date minus seven times the number of completed gestational weeks) and the date of giving birth.

Agreement of vaccination status as determined using the AIR and the PDC was assessed for influenza and pertussis separately, and defined as the proportion of the total number of pregnancies that met one of the following criteria: 1. Vaccinated as per PDC and vaccination recorded on the AIR, 2. Unvaccinated as per PDC and no vaccination record on the AIR, 3. Vaccination status unknown per PDC and no vaccination record on the AIR (as these pregnancies were considered unvaccinated in the PDC-reported coverage estimates).

As reporting to the AIR was mandated during 2021, we present additional details only for pregnancies ending in 2022. We assessed the number of pregnancies during which an influenza vaccine and a pertussis-containing vaccine were co-administered (defined as given on the same day); gestation at vaccination (defined as number of completed weeks gestation on the vaccination date); calendar month at vaccination; and vaccination provider type (community health centre, council, general practice, midwife, nurse practitioner, pharmacy, public hospital, private hospital, commercial, public health unit, state health department, other, unknown, or combined, as appropriate). In the small number of cases where more than one of the same vaccine type was provided during gestation, the most recent dose was used. We also describe agreement between the AIR and PDC for influenza and pertussis vaccination status, separately, by selected characteristics. These include age (<25, 25–29, 30– 34, 35–39 and ≥40 years); socio-economic status (SES) defined using the Index of Relative Socioeconomic Disadvantage (IRSD) in quartiles; remoteness of residence using the Accessibility/Remoteness Index of Australia Plus (ARIA+) categorised as major cities or regional, remote or very remote; and main model of antenatal care (private obstetrician, general practitioner [GP] obstetrician, shared care, public hospital [including public hospital and public hospital high-risk], public midwifery continuity-of-care [including midwifery caseload and team midwifery], and other [including private midwife, combined care, remote area, no formal care and not stated]).

### Ethics Statement

This research was approved by the NSW Population and Health Service Research Ethics Committee (reference number 2021/ETH01116).

## Results

For analyses assessing coverage and agreement over time, after excluding 6,487 pregnancies due to key variables missing, we included 555,553 pregnancies ending between 1 January 2017 and 31 December 2022. These represent 307,758 people with one pregnancy in the study period, 104,659 with two, 11,744 with three, 798 with four or more. There were 852 (0.2%) pregnancies that linked to two or more influenza vaccine doses in the AIR, and 857 (0.2%) had two or more pertussis-containing vaccine doses recorded. For the other analyses, we included only the 89,112 pregnancies that ended in 2022, including 40 people with two pregnancies ending in this year. The characteristics of people with pregnancies ending in 2022 are shown in Table 1.

**Table 1.** Influenza and pertussis vaccination during pregnancy, agreement between the Australian Immunisation Register (AIR) and the New South Wales Perinatal Data Collection (PDC), by selected maternal and pregnancy characteristics, 2022, New South Wales.

|  | All pregnancies ending in 2022, n (col %) | Agreement between AIR and PDC: influenza, n (row %) | Agreement between AIR and PDC: pertussis, n (row %) |
| --- | --- | --- | --- |
| **Total** | 89,112 (100) | 67,323 (75.5) | 50,398 (56.6) |
| **Age at the end of pregnancy (years)** |  |  |  |
| <25 | 9,256 (10.4) | 7,105 (76.8) | 4,691 (50.7) |
| 25–29 | 21,661 (24.3) | 16,441 (75.9) | 11,621 (53.6) |
| 30–34 | 33,440 (37.5) | 25,129 (75.1) | 19,117 (57.2) |
| 35–39 | 20,080 (22.5) | 15,518 (77.3) | 12,150 (60.5) |
| ≥40 | 4,675 (5.2) | 3,520 (75.3) | 2,819 (60.3) |
| **Socioeconomic status, IRSD quintile** |  |  |  |
| 1 (lowest) | 19,894 (22.3) | 15,192 (76.4) | 10,546 (53.0) |
| 2 | 18,222 (20.4) | 13,801 (75.7) | 9,760 (53.6) |
| 3 | 16,880 (18.9) | 12,643 (74.9) | 9,492 (56.2) |
| 4 | 15,049 (16.9) | 11,271 (74.9) | 8,405 (55.9) |
| 5 (highest) | 18,346 (20.6) | 13,906 (75.8) | 11,845 (64.6) |
| Unknown | 721 (0.8) | 510 (70.7) | 350 (48.5) |
| **Remoteness of residence** |  |  |  |
| Major cities | 69,390 (77.9) | 52,422 (75.5) | 40,024 (57.7) |
| Regional, remote or very remote | 19,002 (21.3) | 14,391 (75.7) | 10,024 (52.8) |
| Unknown | 720 (0.8) | 510 (70.8) | 350 (48.6) |
| **Main model of antenatal care** |  |  |  |
| Private obstetrician | 24,224 (27.2) | 17,560 (72.5) | 18,436 (76.1) |
| GP obstetrician | 1,008 (1.1) | 846 (83.9) | 788 (78.2) |
| Shared care | 8,921 (10.0) | 6,830 (76.6) | 5,154 (57.8) |
| Public hospital, conventional | 43,179 (48.5) | 32,789 (75.9) | 20,528 (47.5) |
| Public hospital, midwife-led | 10,619 (11.9) | 8,307 (78.2) | 4,684 (44.1) |
| Other | 1,161 (1.3) | 991 (85.4) | 808 (69.6) |

Abbreviations: AIR = Australian Immunisation Register, GP = General Practitioner, IRSD = Index of Relative Socio-economic Disadvantage, PDC = Perinatal Data Collection.

### AIR and PDC vaccination coverage

PDC-reported coverage peaked in 2020 at 58.8% (52,940/90,091) for influenza and 79.0% (71,169/90,091) for pertussis, before falling to 49.1% (43,744/89,112; 9.4% decrease) for influenza and 77.6% (69,129/89,112; 1.4% decrease) for pertussis in 2022 (Figures 1A and 1B). While available AIR-reported coverage increased over time, it was consistently lower for both vaccines compared to PDC-reported coverage.

**Figure 1.**
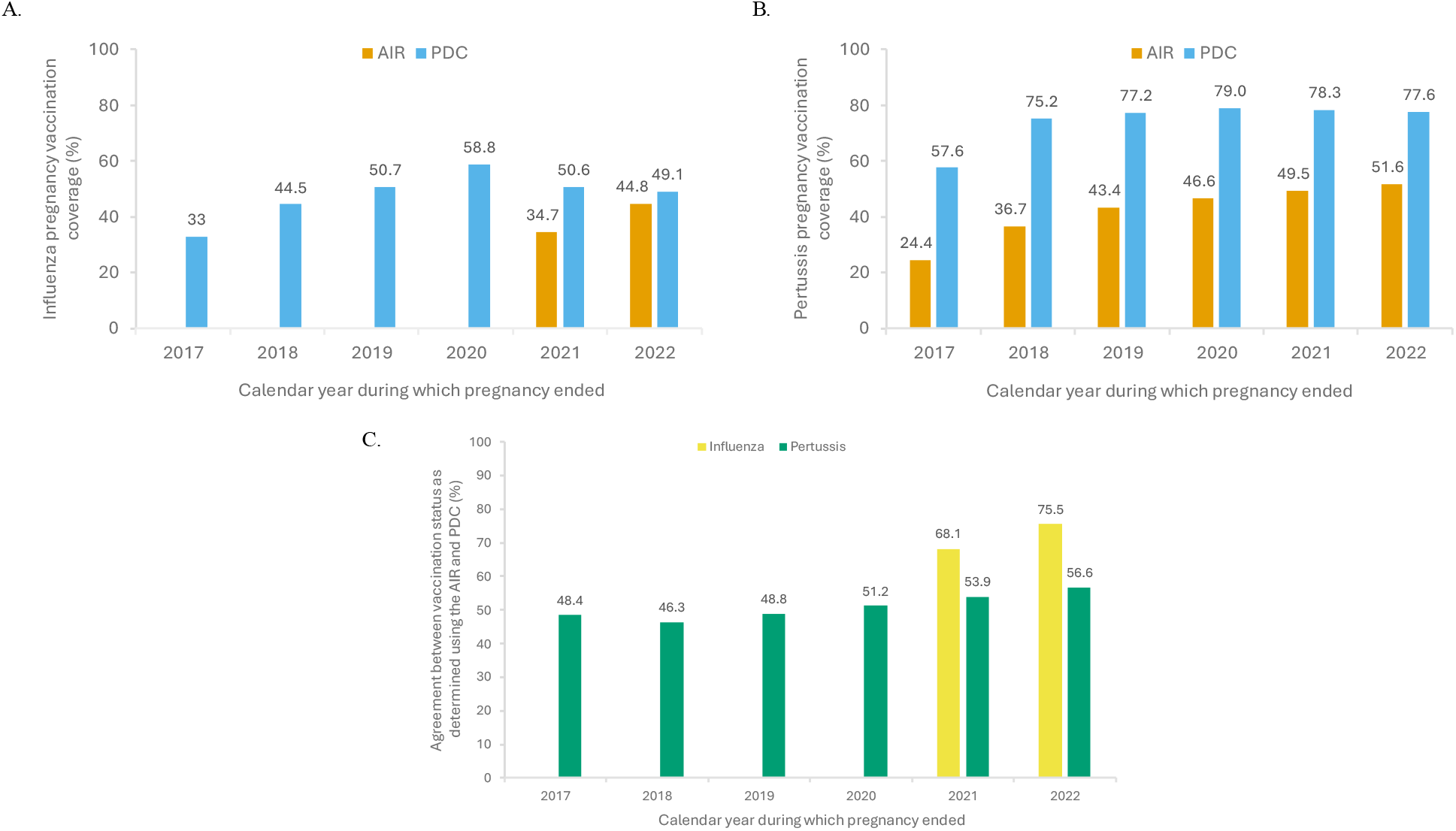
Influenza (A) and pertussis (B) vaccination coverage during pregnancy in the Australian Immunisation Register (AIR) and NSW Perinatal Data Collection (PDC) by year (2017–2022) with AIR-reported vaccination provider type (2022 only), (C) agreement between the two data sources for influenza and pertussis vaccination separately, by year during which the pregnancy ended (2017–2022), New South Wales. Abbreviations: AIR = Australian Immunisation Register, PDC = Perinatal Data Collection.

### Trends in agreement between the AIR and PDC

For both influenza and pertussis, agreement between AIR and PDC-determined vaccination status improved over time (Figure 1C), noting that only two data points were available for influenza. Agreement between AIR and PDC, where available, was consistently higher for influenza than pertussis. In 2022, agreement was 75.5% (67,323/89,112) for influenza and 56.6% (50,398/89,112) for pertussis.

### Vaccination encounters reported to the AIR for pregnancies ending in 2022

In Figure 2 we describe only vaccination encounters that were reported to the AIR in pregnancies ending in 2022. Of 39,935 influenza vaccination encounters reported to the AIR, three-quarters occurred at the GP (n=30,136), followed by pharmacy (n=4,914, 12.3%) and public hospitals (n=2,116, 5.3%), with limited encounters reported by other provider types (Figure 2A). Three-quarters (n=30,582) of influenza vaccinations were administered in April, May and June (Figure 2B). Fewer vaccine encounters occurred in the first 4 weeks of pregnancy and from 36-weeks’ gestation onwards (Figure 2C). Otherwise, distribution was relatively even with small peaks around weeks 14 and 28.

**Figure 2.**
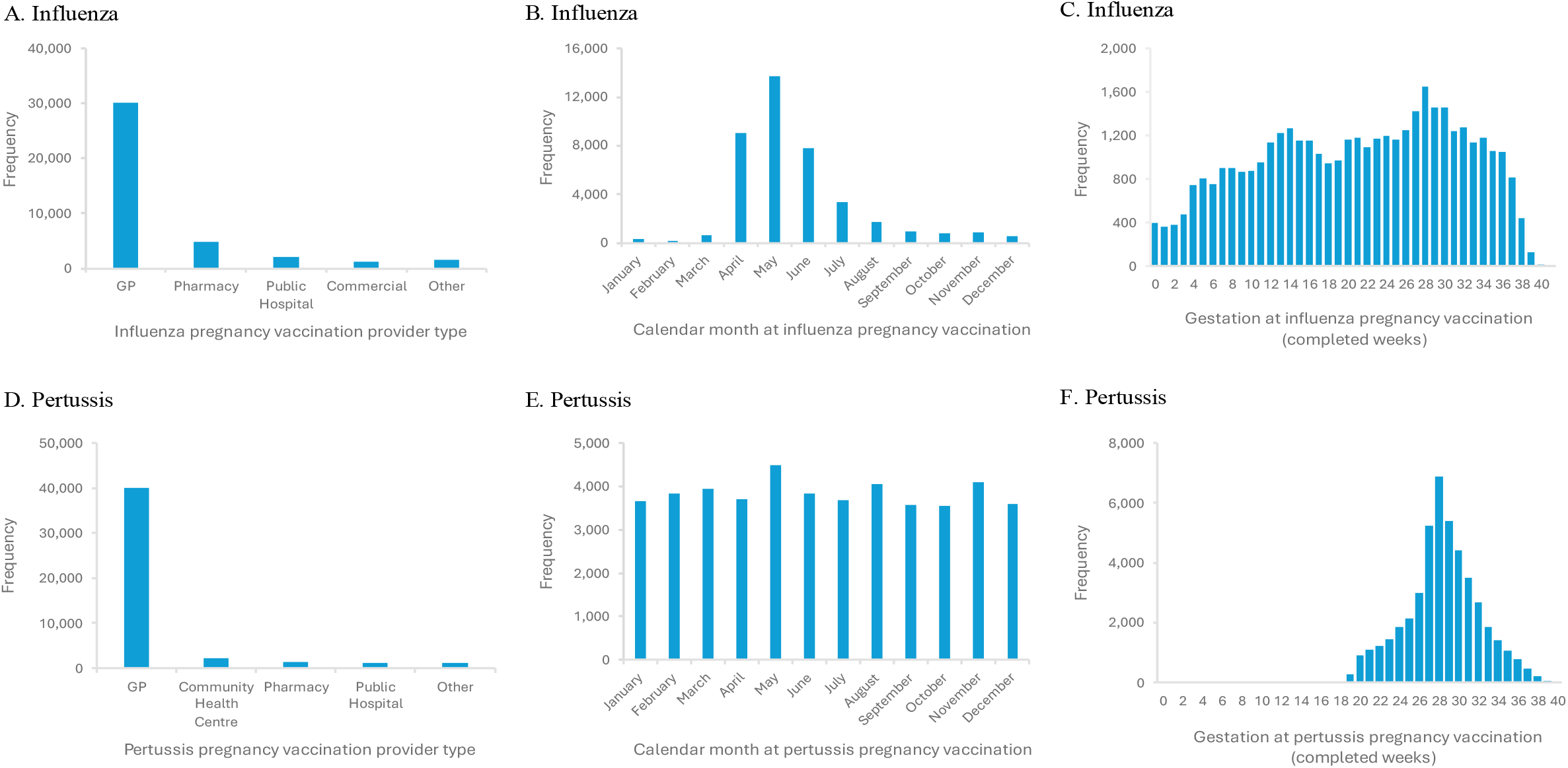
Vaccination provider type, calendar month and gestation at time of influenza (A, B and C) pertussis (D, E and F) vaccinations reported to the Australian Immunisation Register (AIR) in pregnancies ending in 2022, New South Wales. Abbreviation: GP = General Practitioner.

Of 46,009 pertussis vaccination encounters reported to the AIR, 86.9% (n=39,974) were provided in general practice and 5.0% (n=2,288) in community health centres (Figure 2D). Vaccination occurred evenly across calendar months (Figure 2E), from 20-weeks’ gestation onwards with a clear peak at 28-weeks (Figure 2F).

Approximately one-third of pregnancies ending in 2022 (28,725/89,112, 32.2%) had both an influenza and pertussis vaccination reported to the AIR (PDC: 40,665, 45.6%). Of those with both vaccines recorded on the AIR, 27.2% (n=7,804; 8.8% of the total) occurred on the same day. Almost 90% (n=6,975) of these reported co-administration encounters occurred in general practice. The encounters followed the same seasonal pattern as influenza vaccinations, and the same gestational distribution as pertussis vaccinations.

### Agreement between the AIR and PDC for pregnancies ending in 2022

For influenza vaccination, agreement between AIR and PDC varied minimally by assessed characteristics (Table 1). For pertussis vaccination, agreement was better with increasing age and socio-economic status, and was lower for people residing in regional, remote or very remote areas compared to major cities. Agreement between AIR and PDC for pertussis vaccination status was particularly low in the public hospital and midwifery continuity-of-care models (47.5% [20,528/43,179] and 44.1% [4,684/10,619], respectively).

## Discussion

In this assessment of pregnancy vaccinations reported to the AIR and PDC, we found a decrease in PDC-reported coverage since 2020 for both pertussis and particularly influenza. While AIR-reported coverage and agreement between AIR and PDC improved over time, in 2022, pregnancy vaccinations were still substantially underreported to the AIR, particularly pertussis vaccinations. In 2022, most pregnancy vaccinations reported to the AIR were provided in general practice, although this is likely influenced by inconsistent reporting in other provider settings. The timing of pertussis vaccine provision in relation to gestation was in line with clinical guidelines. Influenza vaccines were provided throughout pregnancy, but predominantly in April, May and June. Co-administration of these two vaccines was reported in <10% of pregnancies.

Our study demonstrates the benefits that linkage of the AIR and PDC provides over these data sources separately. We were able to estimate influenza and pertussis pregnancy vaccination coverage using vaccinations reported to the AIR, as linkage with the PDC provided a denominator (i.e. number of pregnancies). The ability to estimate pregnancy vaccination coverage using linked health data is of growing importance as some vaccines that have been recommended to pregnant people are not currently captured on the PDC. A single dose of RSV vaccine between 28-and-36-weeks gestation was routinely recommended in June 2024 (20), and in December 2024 was announced to be included on the NIP from 2025 (21). Monitoring of RSV pregnancy vaccination coverage will depend on accurate reporting to the AIR and linkage with data on the pregnant population.

Through linkage, we could monitor how reporting of pregnancy vaccinations to the AIR improved over time, with reporting in 2021 and 2022 likely influenced by the mandate to report. However, significant underreporting, more so for pertussis, remained despite this mandate. For pregnancy vaccinations, similar to other adult vaccinations (22), underreporting to AIR was much greater than that estimated for childhood vaccinations (23-26).

While reporting to the AIR improved over time, PDC-reported influenza pregnancy vaccination coverage fell by almost 10% between 2020 and 2022, with a smaller drop (1.4%) in pertussis coverage. Decreases in influenza and pertussis pregnancy vaccination coverage since 2020 were also observed in Queensland (12), Victoria (27), and Western Australia (28,29), as well as internationally in the United Kingdom (30,31) and the United States (32). Improving pregnancy vaccination coverage to pre-pandemic levels and above is important with the return of influenza seasons since 2022 and the resurgence of pertussis in 2024 (33). Access to timely, complete, national data is essential to assess both trends in coverage and the impact of interventions to improve vaccine uptake.

Linking the AIR and PDC also provided information about the timing and setting of pregnancy vaccinations. Among pregnancies ending in 2022, timing of vaccination according to gestation was in line with clinical recommendations. While pertussis vaccinations were provided year-round, influenza vaccines were mostly provided in April, May and June, when the new season’s vaccine becomes available, consistent with previous research (6). In Australia, influenza pregnancy vaccination is recommended year-round, within the limits of availability. Strategies to improve influenza vaccine uptake outside of the “vaccination season” should be considered to maximise infant protection. While pregnant people and healthcare providers might be tempted to wait for next season’s influenza vaccine, people giving birth between February and May are less likely to receive any influenza vaccine during pregnancy (10). This impacts those infants born just prior to an influenza season, who are most vulnerable and not yet old enough to be vaccinated themselves during the season.

Given the low instances of co-administration in our data, offering influenza vaccination at the time of pertussis vaccination, if not already received, could be a strategy to increase influenza vaccine uptake and reduce these seasonal gaps, although further work on reasons for low co-administration should also be examined, particularly with a third pregnancy vaccine program (RSV) starting in 2025.

The agreement in vaccination status as determined using the AIR and PDC was predominantly affected by the model of antenatal care, and this was particularly pronounced for pertussis. Reporting of pregnancy vaccinations to the AIR was high in the GP-obstetrician model of care, where the antenatal care provider and vaccination provider are likely to be the same person. In NSW, GPs are often the primary immunisation providers and therefore likely to be aware of recommendations and reporting requirements, with access to automated reporting software in their place of practice. In contrast, agreement between the AIR and PDC for pertussis vaccinations in the public models of care was low. Pregnant people receiving care through the NSW public hospital system may have access to vaccination at the point of antenatal care (e.g. within the antenatal clinic) by providers other than GPs (e.g. midwives, nurse practitioners). We found few vaccinations reported to the AIR that were provided by public hospitals, nurse practitioners or midwives among people birthing in 2022, suggesting that vaccination encounters in these settings are underreported. Possible explanations could be the administrative burden associated with the increasing number of immunisations provided at the point of antenatal care, or access to and functionality of the software used in antenatal clinics to record and transmit vaccination data to the AIR. In the public antenatal care models, there was considerable difference in influenza and pertussis vaccination reporting, which might reflect a difference in provider of these vaccines in this population (e.g. influenza vaccine provided by the GP, pertussis vaccine provided in the antenatal clinic), with different levels of reporting between providers. Improved automated transmission of vaccination records to the AIR from clinical software used in NSW public hospitals is anticipated and, while careful monitoring is required, is expected to improve reporting to the AIR and agreement between the AIR and PDC.

The association between model of antenatal care and reporting to the AIR may also explain the difference in reporting of pertussis vaccinations by age, SES and remoteness of residence, as younger, more socio-economically disadvantaged and regionally or rurally residing people may be more likely to use the public antenatal care models. This is important to consider when interpreting data based on AIR-reported vaccinations, as these pregnant people, as well as providers vaccinating these people, are underrepresented in the data. Age, SES and remoteness of residence are known factors associated with pregnancy vaccination uptake in NSW (10) and other Australian settings (9), thus to optimally utilise the data available in the AIR to guide strategies to improve vaccination uptake, efforts should be made to improve AIR-reporting in these settings.

Study limitations include being unable to assess AIR-reported influenza pregnancy vaccination coverage and agreement between AIR and PDC between 2017 and 2020, as we did not receive AIR data for influenza vaccines provided before 1 January 2021. Thus, the 2021 estimates are likely underestimates, as people giving birth early in 2021 may have received influenza vaccine in 2020. Second, vaccination status in the PDC may not have been validated using (written) medical records resulting in some degree of misclassification. Third, some level of incorrect linkage may have occurred; this is estimated to be <0.5% (18,34).

Lastly, these data represent the NSW population only, and may not be generalisable to other jurisdictions within Australia, as pregnancy vaccination provision and antenatal care models may vary between states and territories.

### Conclusion

Linkage of the AIR to PDC improves our understanding of pregnancy vaccination provision and reporting. Specifically, linkage allows for assessment of pregnancy vaccination timing with regards to calendar time and gestation, vaccination provider types, and vaccination coverage of all vaccines on the NIP, not only those reported to the PDC.

Based on this work and the current policy environment for pregnancy vaccination, we suggest the following priorities:

1. identifying methods that provide timely, national reporting of pregnancy vaccination coverage;
2. monitoring reporting of pregnancy vaccinations to the AIR to ensure improvements continue;
3. supporting the reporting of pregnancy vaccinations in antenatal clinics, and identifying the barriers and facilitators of proactive vaccination strategies;
4. understanding the extent of providers’ knowledge regarding influenza vaccine recommendations, and barriers to co-administration of influenza and pertussis vaccines during pregnancy.

These factors should assist in designing service-specific, evidence-based interventions to improve equitable pregnancy vaccination coverage. This focus is particularly important now with new pregnancy vaccines, such as for RSV, on the horizon.

## Data Availability

The data used in this research are available to researchers on request and subject to approval from the relevant data custodians and ethics committees. The authors do not have permission to share data.

## Acknowledgements

The authors wish to thank NSW Health and Services Australia for providing the data for this study, and the NSW Centre for Health Record Linkage for conducting the data linkage. We would also like to acknowledge Kerry-Ann O’Grady for her valuable review of the manuscript.

